# Assessing the Potential Anti-Neuroinflammatory Effect of Minocycline in Chronic Low Back Pain: Protocol for a randomized, double-blind, placebo-controlled trial

**DOI:** 10.1101/2022.06.22.22276757

**Authors:** Erin J. Morrissey, Zeynab Alshelh, Paulina C. Knight, Atreyi Saha, Minhae Kim, Angel Torrado-Carvajal, Yi Zhang, Robert R. Edwards, Chelsea Pike, Joseph J. Locascio, Vitaly Napadow, Marco L. Loggia

**Affiliations:** MGH/HST Athinoula A. Martinos Center for Biomedical Imaging, Department of Radiology, Massachusetts General Hospital, Harvard Medical School, Charlestown, MA, USA; Medical Image Analysis and Biometry Laboratory, Universidad Rey Juan Carlos, Madrid, Spain; Department of Anesthesia, Critical Care and Pain Medicine, Massachusetts General Hospital, Harvard Medical School, Boston, MA, USA; Department of Anesthesiology, Perioperative and Pain Medicine, Brigham and Women’s Hospital, Harvard Medical School, Boston, MA, USA; Department of Psychiatry, Massachusetts General Hospital, Boston, MA, USA; Harvard Catalyst Biostatistical Consulting Unit, Massachusetts General Hospital, Boston, MA, USA; Department of Physical Medicine and Rehabilitation, Spaulding Rehabilitation Hospital, Harvard Medical School, Boston, MA, USA

## Abstract

**Introduction:** Chronic pain is an extremely prevalent public health issue. However, the underlying mechanisms are poorly understood, thus limiting effective treatment options. Both preclinical studies, and more recent clinical imaging studies, suggest that glia-mediated neuroinflammation may be implicated in chronic pain, and therefore might be a potential treatment target. However, it is currently unknown whether modulating neuroinflammation effectively alleviates pain in humans. This trial tests the hypothesis that minocycline, an FDA-approved tetracycline antibiotic and effective glial cell inhibitor in animals, reduces neuroinflammation and may reduce pain symptoms in humans with chronic low back pain.

**Methods and analysis:** This study is a randomized, double-blind, placebo-controlled clinical trial. Subjects, aged 18-65, with a confirmed diagnosis of chronic (≥ six months) low back pain (cLBP) and a self-reported pain rating of at least four out of ten (for at least half of the days during an average week) are enrolled via written, informed consent. Eligible subjects are randomized to receive a 14-day course of either active drug (minocycline) or placebo. Before and after treatment, subjects are scanned with integrated Positron Emission Tomography/Magnetic Resonance Imaging (PET/MRI) using [^11^C]PBR28, a second-generation radiotracer for the 18 kDa translocator protein (TSPO), which is highly expressed in glial cells and thus a putative marker of neuroinflammation. Pain levels are evaluated via daily surveys, collected seven days prior to the start of medication, and throughout the 14 days of treatment. General linear models are used to assess pain levels and determine the treatment effect on brain (and spinal cord) TSPO signal.

**Ethics and dissemination:** This study was approved by the Massachusetts General Hospital Institutional Review Board (Protocol Number: 2017P000179) and the U.S. Food and Drug Administration (IND Number: 142546). The results of the study will be disseminated via publications in peer-reviewed journals, presentations at conferences globally, and through various media.

**Trial registration number:** ClinicalTrials.gov (NCT03106740)

**Strengths and limitations of this study:**

- This is the first project to image the potential neuro-inflammatory effect of minocycline in a chronic pain condition.
- The use of simultaneous PET/MRI allows us to collect additional, ancillary MRI data that can be used to perform a comprehensive neurophysiological assessment of subjects.
- The sample size (n∼50 per group) is relatively small to detect a clinical response. An additional limitation is the relatively short treatment duration (i.e., 2-weeks).

## INTRODUCTION

Chronic pain is a widespread public health issue^1^ with enormous prevalence. An estimated 35.5% of the general population or 105 million individuals, suffer from chronic pain in the United States^2^. Chronic pain affects both physical and mental functioning, thereby compromising quality of life and affecting astronomical societal and economic costs. These include those directly associated with diagnosing and treating pain, as well as costs resulting from lost work time and reduced productivity^3,4^, and more.

Resources and clinical treatments for managing chronic pain are often ineffective, non-specific, and/or associated with negative side effects. For instance, evidence supporting the long-term effectiveness of opioid drugs in relieving pain and improving functional status is weak^5 6^. Traditional interventions have also been found to be accompanied by a myriad of side effects, including constipation, pruritus (i.e., itching), respiratory depression, nausea, vomiting, hyperalgesia, dizziness, sedation, and importantly, abuse and dependence ^7-10^. Benzodiazepines or other treatments are also often ineffective and associated with negative side effects, such as increased depression^11^. The aforementioned examples stress the importance of achieving a better understanding of the pathophysiological mechanisms underlying chronic pain, to eventually identify alternative and viable treatment options.

Current treatment options for chronic pain are primarily aimed at suppressing neuronal activity within nociceptive pathways of the nervous system; however, it is becoming increasingly clear that non-neuronal cells are involved in the establishment and/or maintenance of clinical pain symptoms. In fact, multiple preclinical studies suggest a central role of glial cells in the nervous system, including microglia and astrocytes, in the etiology of chronic pain^12 13-15^. For instance, both activated microglia and astrocytes can produce cytokines such as IL-6, TNF-α and IL-1β and other pro-inflammatory mediators that are thought to play an essential role in the pathogenesis of chronic pain^16 17^. While evidence of pain-related glial activation was originally observed in the spinal cord, recent preclinical studies show evidence of pain-related glial activation in the brain, including in the rostral ventromedial medulla^18^, the trigeminal nuclear complex^19 20^, the ventral posterolateral nucleus of the thalamus^21 22^, and other brain regions.

Though evidence linking neuroinflammation in human pain is limited, in the past decade our group has shown that subjects with chronic low back pain (cLBP) have increased brain^23-25 26^ and spinal cord^27^ levels of the 18kDa translocator protein (TSPO). TSPO is a five-transmembrane domain protein located primarily at the sites of contact between the outer and inner mitochondrial membranes^28^. While TSPO is ubiquitously expressed throughout the body and across multiple cell types, it is widely used as a marker of neuroinflammation because its expression levels are low in the healthy CNS, but become dramatically upregulated in activated microglia, macrophages and astrocytes during neuroinflammatory responses (at least in preclinical models)^28-30^. Therefore, the TSPO signal elevations we observed in subjects with cLBP suggest that neuroinflammation may occur in chronic pain patients.

The aim of this project is to determine whether the modulation of glial cells is viable and effective as a treatment for cLBP in humans. To do so, we are conducting a clinical trial testing the effect of the FDA-approved tetracycline antibiotic, minocycline, on neuroinflammation and pain symptoms. In pre-clinical models, minocycline is commonly used as a glial inhibitor, including in pain models^22 31-34^. While minocycline is mainly used to treat bacterial infections in humans, a recent study showed that it induced a small but statistically significant reduction in pain^35^, although it remains unknown whether this effect might be mediated by an effect on glia. To measure the effect of minocycline on TSPO signal, we are using PET/MRI with [^11^C]PBR28, a second-generation TSPO radioligand^36 37^.

## TRIAL OBJECTIVES

The primary purpose of this study is to evaluate whether minocycline exerts anti-neuroinflammatory effects in the brains of subjects with cLBP. Secondary outcomes include spinal cord inflammation and pain ratings. Additionally, we will test exploratory hypotheses that higher pre-treatment neuroinflammatory signal predicts larger behavioral and imaging responses to minocycline. By assessing the effect of minocycline on glial activation and pain, and the role of inflammation as a predictor of response to minocycline, we will develop a better understanding of the mechanisms of chronic pain, and the role of neuroinflammation as a potential target for treatment.

## METHODS AND ANALYSIS

### Trial design

This is a Phase II, double-blind, placebo-controlled study. A total of ∼50 subjects will complete participation in the study, randomized to receive either Minocycline 100mg (N∼25) or placebo (N∼25). Subjects are considered to have completed the study once they have been enrolled (via written, informed consent), scanned with integrated Positron Emission Tomography/Magnetic Resonance Imaging (PET/MRI) prior to treatment, and re-scanned with PET/MRI upon conclusion of the trial period (Figure 1). Low back pain at baseline is determined using daily surveys administered for seven days prior to the start of medication, while treatment effect on pain is assessed via daily surveys administered during the treatment period. This trial is being conducted at Athinoula A. Martinos Center for Biomedical Imaging in the United States. Recruitment began in 2017 and is expected to conclude in Summer 2022.

**Figure 1.**
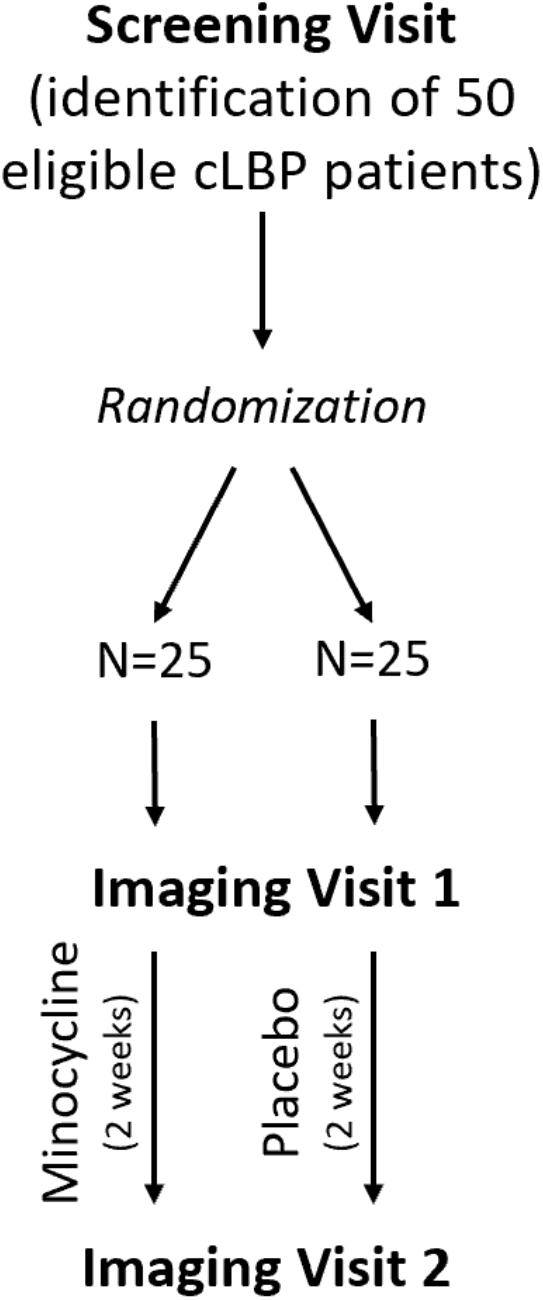
Study schema.

### Population

We will recruit approximately 50 subjects with cLBP, aged 18 to 75 through community advertising, clinical research databases, and social media platforms. In order to be considered eligible for this study, subjects must have cLBP for at least six months, supported by medical records confirming their diagnosis. CLBP must be ongoing for at least 6 months prior to enrollment, with pain averaging at least four out of 10 (on a 0-10 scale) and present for at least half of the days during a typical week. Subjects must not undergo any new interventional pain procedures between the two scans, and any pain treatment (pharmacological or otherwise) must be stable for at least four weeks prior to enrollment. Exclusion criteria include contraindications to study procedures-including to MRI and PET scanning or minocycline administration. Additionally, a detailed list of all inclusion and exclusion criteria are outlined in Table 1.

**Table 1.**
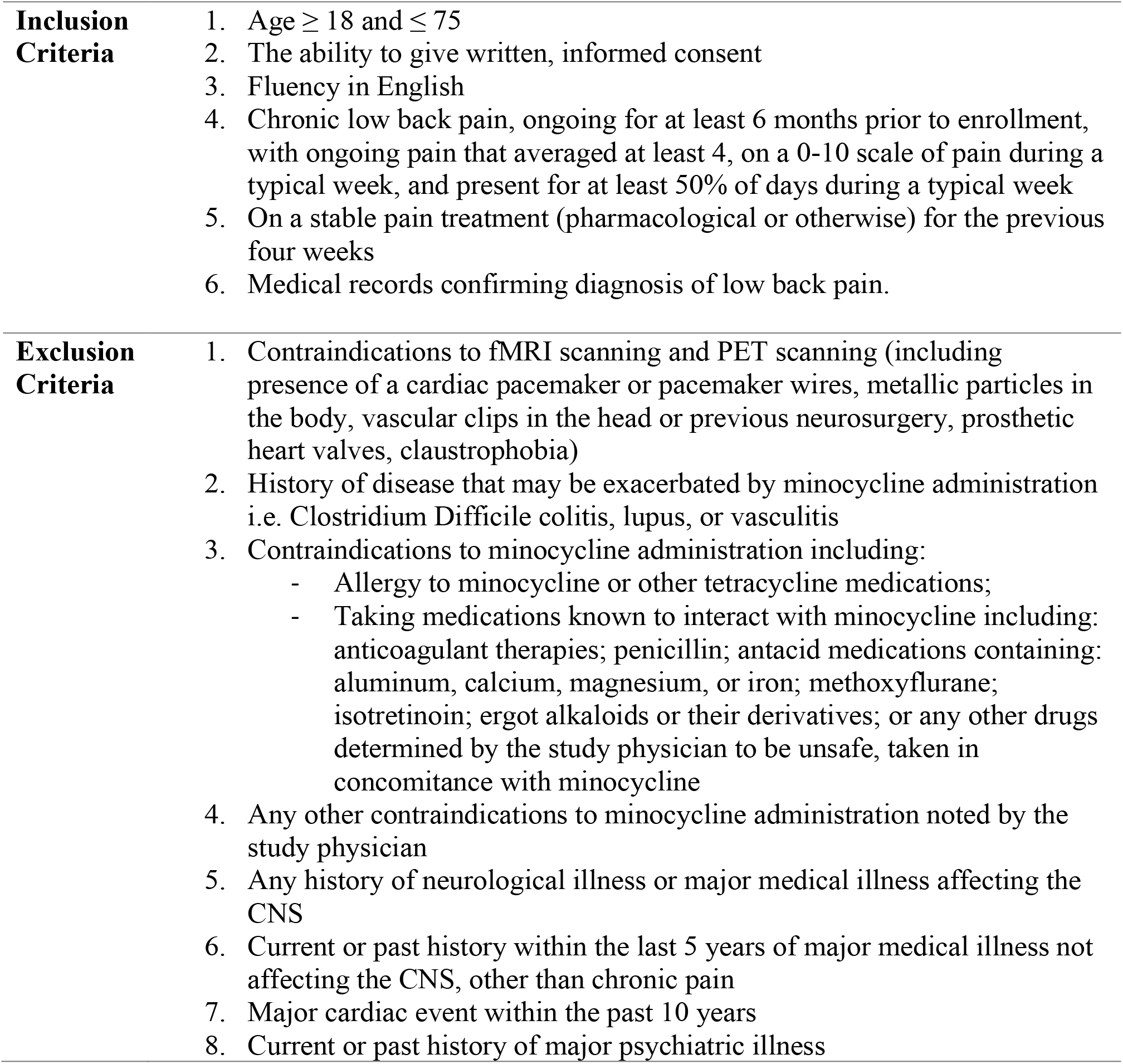
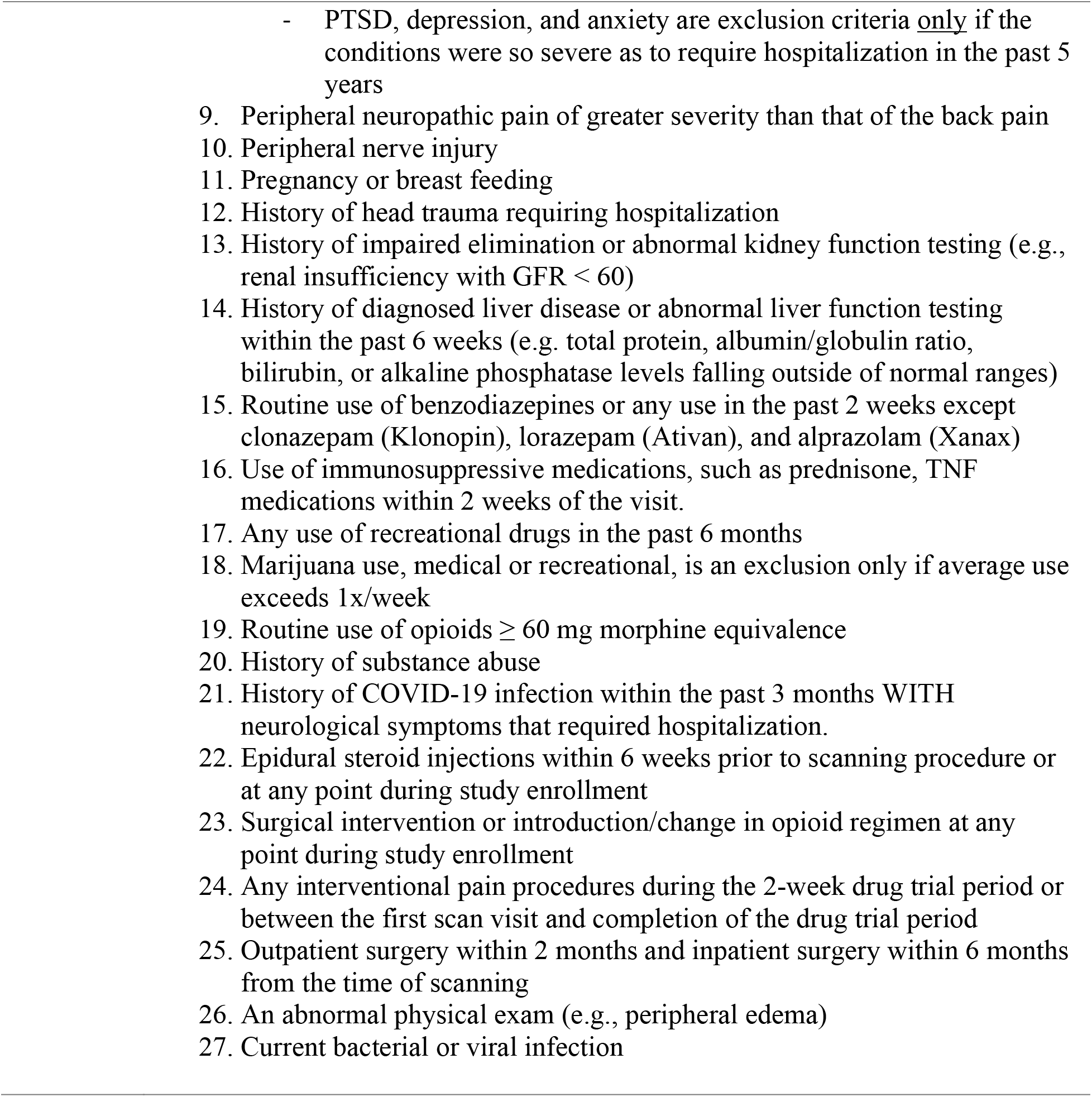
Detailed list of all inclusion and exclusion criteria

### Intervention and randomization

Following enrollment, subjects are randomized in a 1:1 ratio to receive either placebo or minocycline 100mg once daily for two weeks. A randomization number is assigned to each subject via a computerized random number generator to determine treatment group. The Clinical Trials Pharmacy at Massachusetts General Hospital is maintaining subject treatment assignments (i.e., minocycline or placebo) for identification upon completion of the trial. Therefore, subjects and study staff are blinded to group assignment. Study procedures are supplemental to routine clinical care and maintained throughout participation. The study drug is added to subjects’ current medication regimen so there are no adjustments to current medications and dosage. The Jackson Compounding Center at Massachusetts General Hospital prepares the minocycline and placebo, encapsulated in identical opaque capsules. Minocycline and placebo are stored at a controlled room temperature out of direct sunlight. To track treatment compliance, both minocycline and placebo have 25mg of riboflavin (vitamin B2) added. Riboflavin acts as a tracer and allows study staff to conduct urine testing using a UV light to ensure that subjects are taking the medication as instructed.

### Investigational product

Minocycline is an FDA-approved antibiotic commonly used to treat infections and is generally considered a safe drug^38^. The most noted side effects are dizziness, drowsiness, headache, photosensitivity, and gastrointestinal disturbance. Dosage and administration protocol for minocycline HCl (i.e., 100mg capsules once daily, by mouth for 2 weeks) was informed by Vanelderen et al., who demonstrated a reduction in pain in a smaller number of subjects with subacute lumbar radiculopathy^35^.

### Screening visit

Subjects deemed preliminarily eligible are invited for a screening visit (either in-person or via HIPAA compliant video call) during which a signed informed consent form is obtained, the procedures involved in the experiment are explained, and several validated assessments are administered (Table 2).

**Table 2.** Questionnaires and assessments administered during each visit.

**Behavioral Session (Visit 1)**
| **Assessment** | **Description** |
| --- | --- |
| *Social Provisions Scale (SPS)* | The social provisions scale measures the social support provided by the subject's current relationships. |
| *Fibromyalgia Survey Questionnaire (FSQ)* | The Fibromyalgia Survey Questionnaire is a 5-item questionnaire that is aimed at assessing the presence of fibromyalgia, or pain centralization, sub-clinically in people who do not meet the fibromyalgia criteria. |
| *The Pain Catastrophizing Scale (PCS)* | It is a 13-item self-report scale which measures pain-related Rumination, Magnification and Helplessness. |
| *Hospital Anxiety and Depression Scale (HADS)* | The HADS is a 14-item self-report survey designed for populations with medical illness<sup>46</sup>. It does not include somatic symptoms, such as fatigue and sleeplessness, which may otherwise be attributable to pain. It asks subjects to rate depression and anxiety symptom over the past week on a 4-point Likert scale. It has been validated in several medical illness populations and has |
|  | been used extensively in subjects with chronic pain. |
| *Adverse Childhood Experiences Scale (ACE)* | The ACE scale asks about 10 common adverse events/experiences that affect development such as childhood maltreatment, parental divorce, and parental incarceration. Increased ACE scores have been associated with negative health outcomes such as shortened life span, mental illness, and chronic illness. |
| *Brief Trauma Questionnaire (BTQ)* | The BTQ will be used to determine whether an individual meets Criterion A1 for traumatic exposure according to the DSM-V PTSD diagnosis. It is a brief self-report questionnaire designed to assess 10 traumatic events including physical assault, car accidents, natural disasters, and unwanted sexual contact. It is derived from the *Brief Trauma Interview*. |

**Imaging Sessions (Visits 2 & 3)**
| **Assessment** | **Description** |
| --- | --- |
| *Patient Reported Outcomes Measurement Information System (PROMIS-29) questionnaire* | The PROMIS-29 is a 29-item self-report measure assessing physical, mental, and social health. |
| *Brief Pain Inventory (BPI)* | The BPI is a 15-item questionnaire assessing pain location, and 0–10 ratings of pain intensity, relief, quality, pain-related quality of life, and function. It has been validated in cancer and noncancer pain conditions. |
| *Snaith-Hamilton Pleasure Scale (SHAPS)* | The Snaith-Hamilton survey is a 14-item questionnaire designed to estimate the degree to which an individual is able to experience pleasure, or the anticipation of pleasure. The questionnaire assesses four domains of hedonic experience: interest/pastimes, social interaction, sensory experience, and food/drink. All items relate to daily experiences encountered by most people. |
| *PainDETECT* | The PainDETECT is a screening questionnaire used to estimate the likelihood of a neuropathic component in chronic pain. |
| *Beck Depression Inventory (BDI)* | The 21-item BDI has shown good sensitivity and specificity for major depression in subjects with chronic pain. |
| *Rey-Osterrieth Complex Figure (ROCF)* | Commonly used neuropsychological assessment tool. This is a visuo-constructional ability and visual memory test comprised of a copying component and recall component. |
| *Trail Making Test, Part B* | A timed neurocognitive tool to test cognition via connecting letters and numbers quickly and accurately. Time to complete the test and number of mistakes combine to measure neurocognition. |
| *Patient Global Impression of Change (Visit 3 Only)* | A questionnaire to assess the overall change perceived by the subject during treatment. |

A trained medical professional (e.g., MD or NP) performs a formal physical examination, including a modified Allen’s test to evaluate the safety of the arterial line placement (for patients that consent to this procedure). The provider verbally collects subjects’ medical history and current medication list to confirm study eligibility.

Next, subjects are genotyped for the Ala147Thr TSPO polymorphism, which predicts high (Ala/Ala), mixed (Ala/Thr) or low (Thr/Thr) affinity binding to the radioligand used for PET imaging^39 40^, via collection of either up to 10 mL peripheral blood or a saliva sample. Subjects exhibiting the low affinity binding genotype are not considered eligible for the [^11^C]PBR28 scan.

To comply with public health efforts to address COVID-19, several study procedures typically performed in-person (e.g., questionnaire administration) are performed during virtual visits, as necessary. Such visits are conducted via Institutional Review Board (IRB) approved and secure online platforms (i.e., Microsoft Teams, Doximity Dialer for communication; REDCap for FDA compliant e-consent).

### Imaging visit

Subjects participate in two imaging visits: before and as soon as possible (i.e., 0-2 weeks) after minocycline or placebo treatment. At the beginning of each scan session, subjects complete screening checklists for PET/MRI scanning to confirm safety and compatibility with the equipment and procedures. An intravenous (IV) catheter is placed in the subject’s antecubital vein of the left or right arm, for the purpose of radiotracer injection as well as to draw blood to confirm a negative pregnancy test (i.e., serum hCG levels) for female subjects of childbearing age, and for additional, exploratory analyses (e.g., of circulating level of inflammatory markers and antibody serology testing for prior SARS-CoV-2 infections). An arterial line is placed in the radial artery in the opposing arm of IV placement under local anesthesia using a sterile technique, for subjects who consent to this procedure. The arterial line enables blood sampling at various times during imaging for radiotracer quantification. Additionally, before subjects undergo imaging, we perform a urine test to screen for use of opioids and illicit drugs (e.g., amphetamine, barbiturates, cocaine, and marijuana), as well as to monitor urine riboflavin levels to aid in the assessment of trial compliance. Riboflavin is measured using a UV light to detect post-treatment changes in riboflavin excreted through the urine, thereby measuring compliance with the prescribed treatment regimen.

Brain PET/MRI data are acquired for approximately 90 minutes post-injection. Images are acquired through integrated PET/MRI using [^11^C]PBR28 produced by the radiopharmacy facility at the A. A. Martinos Center for Biomedical Imaging. Up to 15 mCi [^11^C]PBR28 (i.e., ∼3.7mSv) is injected intravenously with a slow bolus over a 30-60 second period by a licensed nuclear medicine technologist. Safety standards approved by the Radiation Safety Committee for the use of radioligands are strictly followed. Subjects are instructed to remain still, with eyes open, for the total duration of the scans. Before, during, and after the scan, current severity of low back pain and anxiety are assessed (expressed by subjects either verbally or using a button box connected to a computer). Vital signs are collected at the beginning and end of each scan visit to assess for any adverse events in relation to [^11^C]PBR28 administration.

Additionally, functional MRI (fMRI) data is simultaneously collected to evaluate the effect of the treatment on resting-state functional connectivity, activation of reward circuitry (as assessed using the Monetary Incentive Delay (MID) task^41^; for methods see ^42^), brain activation in response to somatosensory stimulation (see ^25^) and other ancillary brain measures, in exploratory analyses. For subjects who consent to this procedure, electrical stimulation is applied to the back and legs, using electrodes connected to a transcutaneous electrical nerve stimulator, to assess the relationship between brain activation in response to innocuous and tolerable painful stimuli and neuroinflammation, as well as to functionally define the S1 brain representation of body regions affected by low back pain (back and legs)^43^. Electrodes are placed on the left and right of the fourth lumbar spine vertebra, and on the lateral and medial sections of both knees. Subjects undergo two separate stimulation runs, each lasting 5 minutes and 38 seconds. In each run, subjects receive five electric current stimuli at each of the three body regions, i.e., back, right leg and left leg in pseudo-randomized order. Each electrical stimulus is applied for two seconds at 35 Hz, at either 5mA (first run) or 12mA (second run). Stimuli are delivered using a TENS unit (Empi 300pv electrotherapy system) controlled with an in-house script using LabVIEW 16, Austin, Texas. Each stimulus is delivered 4-8s (jittered) after a visual anticipatory cue, indicating the body part about to be stimulated (indicated by the words “Back” “Right leg”, or “Left leg” projected in black onto a white background). Visual stimuli are presented using E-Prime (version 2.0). At the completion of each run, subjects are asked to rate the average pain intensity from 0-100 (0 indicating no pain and 100 indicating worst pain tolerable) at each site.

During the scan we also perform single-voxel Magnetic Resonance Spectroscopy (MRS), allowing us to determine the effect of the treatment on thalamic levels of various neurometabolites, such as of N-acetylaspartate (NAA; a marker of neuronal integrity), myo-inositol (mIns; a glia marker), and choline (Cho; a cell membrane metabolism and cellular turnover marker often linked to neuroinflammatory processes) (see ^44^).

After the brain scan (i.e., at 90 minutest post-injection), an optional ∼20-minute thoracolumbar spinal cord scan is collected to evaluate the hypothesis that minocycline reduces TSPO signal in the most caudal segments of the spinal cord (at or below T11, as these segments have demonstrated TSPO signal increase in our prior study of subjects with lumbar radiculopathy, compared to healthy volunteers^27^).

During each imaging visit, subjects are asked to complete additional questionnaires to assess various domains (e.g., pain, depression, catastrophizing, etc.), as well as neuropsychological assessments (to test cognitive performance) (see Table 2). Additionally, questions are administered prior to the first scan to assess the participants’ expectation of treatment effect (i.e. “How much do you expect the active medication (minocycline) will relieve your back pain?” and “How likely do you think you will receive the active medication (minocycline)?), and the Patient Global Impression of Change is administered on the day of the second scan, to assess the overall change perceived by the subject during treatment^45^.

### Daily surveys

Throughout the study, daily surveys are administered to assess pain symptoms of subjects with cLBP, and to confirm medication compliance. Surveys are administered for a total of 21 days (i.e., seven days prior to the start of treatment/placebo, and until the end of the 14-day treatment period), online or by phone (via StudyTrax).

### Data analysis

Our primary metric for brain [^11^C]PBR28 signal quantification will be standardized uptake value ratio (SUVR), using the whole brain signal as the normalizing factor, as described in prior work^47-49^. In subjects with arterial blood data available, we will compute distribution volume (V_T_) and ratio of distribution volume (DVR), which will be used as secondary outcome measures and to support the use of SUVR as an outcome metric.

We will perform paired analyses in subjects receiving minocycline or placebo (pre vs. post treatment) and test for a treatment*time interaction in order to assess the effect of minocycline on PET signal (i.e., the primary outcome), as well as pain measures (i.e., secondary outcome). Pain measures will comprise of the difference between the averaged seven pain ratings from the daily surveys pre-treatment and the averaged last seven days of treatment. The difference between these two averages will be compared across groups. In addition, we will analyze the full granular 21 timepoint pain measure data with mixed effects longitudinal models in which linear and quadratic effects of time interacting with Treatment Group will be assessed. It is possible that a significant and meaningful time-related pattern of differences between the Treatment Groups will be detected that was hidden in the analysis above. The Pre-versus Post-Intervention Epoch distinction will also be included as a time-varying binary predictor in the model as well as a 3-way interaction of this with the incremental Time variable and Treatment Group. Other relevant subject-level and time-varying predictors and covariates will be included as warranted substantively or statistically. The effects above would be fixed, whereas subjects would be a random effect possibly interacting with time components.

We will perform a comparison of the imaging data between groups, including TSPO genotype and, if applicable, with age and sex as covariates of no substantive interest. Secondary regression analyses will be performed to evaluate the association between imaging and behavioral data (e.g., questionnaire scores, pain duration, ancillary imaging data). These analyses will be performed as both thalamus-focused ROI and voxelwise approaches, using methods similar or identical to those we have previously used^47^. All behavioral, demographic and ROI data will be analyzed using Statistica 10.0 (StatSoft Inc., USA), and an alpha level of 0.05.

Finally, in addition to the aforementioned analyses, the effect of prior exposure to coronavirus (as detected by the presence of antibodies to SARS-CoV-2) will be evaluated in both ROI and voxelwise analyses. While we explicitly exclude subjects with recent history of severe COVID-19 infection (to avoid the potentially confounding acute/subacute effects on our main analyses on the role of neuroinflammation), identifying subjects who are positive to the antibodies might allow us to test the exploratory hypothesis that prior exposure to the coronavirus can lead to neuroinflammation even without acute COVID-19 symptoms^50^.

#### Box 1. Outcome measures

**Primary Outcome Measures**

1. TSPO signal in the brain (as measured with [^11^C]PBR28 PET)

**Secondary Outcome Measures**

1. Daily pain ratings (as measured by the Brief Pain Inventory, BPI^51^ severity items, daily during treatment/placebo)
2. TSPO signal in the spinal cord (as measured with [^11^C]PBR28 PET)

**Exploratory Outcome Measures**

1. Anxiety and depression symptoms (Hospital Anxiety and Depression Scale, HADS^52^; Beck Depression Inventory, BDI; Snaith-Hamilton Pleasure Scale, SHAPS^53^)
2. Pain catastrophizing (Pain Catastrophizing Scale, PCS^54^)
3. Probability of neuropathic pain component (PainDETECT^55^)
4. Exposure to traumatic events (Brief Trauma Questionnaire, BTQ^56 57^)
5. Negative health outcome associated with adverse events in childhood (Adverse Childhood Experiences Scale, ACE^58^)
6. Social support provided by relationships (Social Provisions Scale, SPS^59^)
7. Degree of nociplastic pain (Fibromyalgia Survey Questionnaire, FSQ^60^)
8. Health-related quality of life (Patient Reported Outcomes Measurement Information System-29, PROMIS-29^61^)
9. Treatment expectancy and Patient Global Impression of Change^45^
10. Brain response to acute pain via electrical stimulation
11. Striatal activation in response to a reward task (Monetary Incentive Delay Task, MID^41^)
12. Cognitive flexibility, working memory, and visual and executive functioning (Trail Making Test, Part B^62^; Rey-Osterrieth Complex Figure^63 64^)
13. Ancillary neuroimaging measures: BOLD fMRI, Diffusion-Tensor Imaging, and Magnetic Resonance Spectroscopy (MRS)

### Power analysis

Because relevant [^11^C]PBR28 PET imaging data were unavailable at the time of trial initiation to inform a power analysis of a treatment effect, we ran a power analysis of treatment effect on expected change in pain ratings, based on a recent clinical trial using minocycline in subjects with back pain^35^. We computed the sample size required for a mixed effects between-subject (Treatment Group: Minocycline/Placebo) and within-subject (Time: Pretest/Posttest) repeated measures ANOVA design. Alpha was set at 0.05 (two tailed test). We assumed a within group/timepoint standard deviation of ∼2 and an estimated test-retest correlation of r∼0.65, based on previous data^35^,^65^. We expressed our anticipated effect size in terms of a pattern of means that we felt realistically mirrored the minimally clinically important effect to which we wished to be sensitive. Specifically, for each group we assumed a mean pain rating of 5 at pretest, but with the placebo mean declining to 4.9 at posttest due to a minor placebo effect whereas the minocycline group would decline to 3.5 due to the drug (a clinically meaningful decline). For these specifications and 80% power to detect the effect of primary interest, i.e., the interaction of Treatment Group X Time, the total sample size required for the study was computed to be 48 (i.e., 24 per group).

### Interim analysis

Results pertaining to the study drug (minocycline) will be determined only once data collection is complete to protect the integrity of the double-blind study design. However, analysis on pre-treatment data are being conducted (for instance, to compare neuroinflammatory data between cLBP of different etiology; see ^43^).

### Unblinding

Subjects and study staff are blinded to group assignment to protect the integrity of the double-blind study design. Unblinding will only occur if a participant experiences an adverse event related to the study procedures and deemed clinically necessary by the study physician (Y.Z.). In this case, the clinical trials pharmacy will facilitate the unblinding of the subject’s treatment allocation.

### Data Safety and Monitoring Board

Adverse events and outcome monitoring are reported to the Mass General Brigham (MGB) IRB and to the FDA, when appropriate. The Principal Investigator (PI) is responsible for communicating with the IRB about any adverse event. Additionally, the MGH Human Research Committee and Radiation Safety Committee is involved as any additional potential risks arise. Should the rate and severity of adverse events result in an unacceptable risk/benefit ratio, the study will be terminated.

All subjects who undergo an arterial line placement are contacted by a member of the study team up to 48 hours following catheter removal to confirm no adverse side effects. Vital signs are also recorded for all subjects before and after [^11^C]PBR28 injection to monitor for any adverse events related to radioligand administration. Additionally, the Siemens PET/MRI scanners used in this study have a built-in self-monitoring system that automatically shut the scanner off if parameters (e.g., scanner bore temperature) exceed safe levels. To further protect the safety of the subjects, the total possible maximum radiation received by the subjects participating in two scans (∼7.4mSv) is significantly less than the maximum allowed annual whole-body dose approved by the U.S. Food and Drug Administration and MGB IRB at 5 REM, i.e., 50mSv^66^. Subjects are informed of all potential adverse effects of minocycline and are asked to immediately contact the study physician (Y.Z.) if they experience side-effects pertaining to the drug.

### Confidentiality

Study staff adheres to the confidentiality requirements set by the MGB Human Research Committee. Data is password-protected and/or stored in secure areas. Stored samples are labeled with deidentified information that does not allow for association with any subject.

## Data Availability

All data and materials used in the analyses may be provided by Marco L. Loggia and Massachusetts General Hospital, pending scientific review and a completed data use agreement/material transfer agreement beginning one year after publication of the results. Requests for all materials should be submitted to Marco L. Loggia at.

## ETHICS AND DISSEMINATION

This study was approved by the Massachusetts General Hospital Institutional Review Board and the U.S. Food and Drug Administration. The proposed study is regularly monitored for safety, and all adverse events and adverse treatment outcomes are reviewed and reported within approved timelines. The PI also routinely monitors and assures the validity and integrity of collected data and assures adherence to the IRB-approved protocol.

The results of the study will be disseminated via publications in peer-reviewed journals, presentations at national and international conferences, and through social and news media. Protocols and results of the study will also be available for public review on ClinicalTrials.gov upon completion of data collection. Neither subjects nor the public were involved in the development, design, or conduct of this study.

## Authors’ contributions

EJM drafted the manuscript and coordinated the project. MLL conceptualized the research question and trial design, obtained funding, and led the project. ZA, AT-C, RRE and VN advised on study procedures. PCK, AS, and MK coordinated the project. YZ is the study physician responsible for obtaining formed consent, performing clinical characterization of cLBP, and overseeing trial safety. CP assisted with manuscript revision. JL advised statistical aspects of the protocol. All authors have approved the final manuscript.

## Subject consent for publication

Obtained.

## Ethics approval

Massachusetts General Brigham IRB, Protocol Number 2017P000179.

